# Data-Driven Patterns in Protective Effects of Ibuprofen and Ketorolac on Hospitalized Covid-19 Patients

**DOI:** 10.1101/2021.06.15.21258991

**Authors:** Benjamin J. Lengerich, Rich Caruana, Yin Aphinayanaphongs

## Abstract

The impact of nonsteroidal anti-inflammatory drugs (NSAIDs) on patients with Covid-19 has been unclear. A major reason for this uncertainty is the confounding between treatments, patient comorbidities, and illness severity. Here, we perform an observational analysis of over 3000 patients hospitalized for Covid-19 in a New York hospital system to identify the relationship between in-patient treatment with Ibuprofen or Ketorolac and mortality. Our analysis finds evidence consitent with a protective effect for Ibuprofen and Ketorolac, with evidence stronger for a protective effect of Ketorolac than for a protective effect of Ibuprofen.

---

The impact of nonsteroidal anti-inflammatory drugs (NSAIDs) on patients with Covid-19 has been unclear. Initial guidance was to avoid prescription of NSAIDs to Covid-19 patients [1]. However, more recent meta-analyses have suggested that there is no compelling evidence of a harmful effect of ibuprofen on Covid-19 patients [2], [3], [4]. It is difficult to identify the effect of NSAIDs due to confounding; beyond association with long-term comorbidities, NSAID usage in Covid-19 patients may also be caused by more severe cases of Covid-19 which could manifest with inflammatory symptoms [5]. However, other analysis has found no association between ibuprofen and clinical outcomes, even without deconfounding [6].

Moreover, a few observational analyses have indicated that a protective effect from outpatient NSAIDs [7] or ibuprofen ^1^ appears after correcting for underlying patient risk factors. Finally, there is some evidence of protective effects of NSAIDs on patients with respiratory illnesses [8], although similar clinical trials have not been completed for Covid-19.

In this setting, we seek to use machine learning tools to estimate patient risk at time of admission from patient lab values and then estimate the additive effect of Ibuprofen and Ketorolac. Our outcome is in-hospital mortality. We consider only treatments within 24 hours of hospital admission; this captures a total of 97 patients who were treated with either Ibuprofen or Ketorolac. Our control condition is not receiving either NSAID in the first 24 hours. Our analysis includes deidentified records from 3108 patients hospitalized for Covid-19. This cohort includes patients hospitalized from March to August 2020 with an average mortality rate of 18.1%. In this dataset, mortality rate peaked over 25% and decreased to less than 5% for patients admitted in August 2020.

Treatment assignment is not random and has changed over time with more glucocorticoid prescriptions at later dates [9]. To correct for this and other confounding, we use a two-stage machine learning procedure to estimate the adjusted risk difference (ARD) and adjusted risk ratio (ARR) [10, 11], which can be interpreted as an odds ratio after correcting for patient mortality risk at admission. Our two-stage procedure works as follows: first we take half of the control patients and train a generalized additive model (GAM) [12, 13] to predict mortality from patient features at admission. Because medications can affect patient lab values, we use only initial lab tests which are taken before in-patient medications are administered. In the treated patients (held out from model estimation), we compute the excess risk after accounting for the model predictions of patient risk. To compute standard error, we bootstrap the training sample. The baseline mortality risk model achieves AUC 0.91 on held-out patients. See S3 for details of model construction.

Results of this analysis are shown in Table 1. The observed mortality rate of patients treated with Ibuprofen or Ketorolac is lower than would be expected based on patient risk factors (comorbidities, demographics, and lab tests) at admission. For Ketorolac, the evidence is sufficient to reject a null hypothesis of no effect at p=0.05.

**Table 1:** Observed and expected mortality rates for patients treated with Ibuprofen or Ketorolac.

| Drug | N | Obs. Mort. (%) | Exp. Mort (%) | ARD (95% CI) | ARR (95% CI) |
| --- | --- | --- | --- | --- | --- |
| Ibuprofen | 55 | 5.17 ± 3.06 | 7.93 ± 4.77 | -2.82(-6.41, 1.80) | 0.65(0.45, 1.54) |
| Ketorolac | 50 | 2.46 ± 1.98 | 6.78 ± 3.81 | -4.29(-7.95, -1.08) | 0.36(0.24, 0.69) |
| Ibuprofen or Ketorolac | 97 | 4.42 ± 2.02 | 7.97 ± 3.21 | -3.50(-6.51, -0.74) | 0.56(0.40, 0.86) |
| Outpatient Aspirin | 197 | 20.73 ± 2.89 | 26.48 ± 4.58 | -5.75(-10.51, -1.32) | 0.78(0.66, 0.94) |

These results add to the growing body of evidence that NSAIDs, and in particular Ibuprofen or Ketorolac, may not have detrimental effects on patients hospitalized with Covid-19. While these medications were correlated with a younger subset of the hospitalized patients (Figure. S3) and later treatment dates (Figure. S2), the protective effects remain after correcting for confounding. This suggests that further study should be performed to understand any potential benefits of Ibuprofen and Ketorolac on patients hospitalized with Covid-19. Our study comes with all caveats of observational analyses and we suggest randomized control trials to study treatment protocols.

## Data Availability

The data include anonymized medical records, which are not publicly available.

## Footnotes

1 https://ehrnprd.blob.core.windows.net/wordpress/pdfs/Ibuprofen-and-COVID-19-Severity.pdf

