## Supplement for "Data-Driven Patterns in Protective Effects of Ibuprofen and Ketorolac on Hospitalized Covid-19 Patients"

### S1 Dataset Construction

Our dataset consists of 11080 total hospitalized patients who have lab-confirmed cases of Covid-19. To filter out patients who were hospitalized for reasons other than Covid-19, we excluded patients who have indicators of (1) pregnancy: outpatient prenatal vitamins, in-patient oxytocics, folic acid preparations; or (2) scheduled surgery: urinary tract radiopaque diagnostics, laxatives, general anesthetics, antiemetic/antivertigo agents, or antiparasitics. We also require that the patients have recorded temperature, age, BMI, and Admission Day. Finally, we remove patients who died within six hours of admission.

To correct for patient risk confounding, we observe pre-admission features including demographics, comorbidities, outpatient medications, initial in-patient vitals, and initial in-patient lab tests. We exclude any measurement taken within 24 hours of the patient mortality. The 45 total features are listed below.

- Demographics/Vitals:
  - Age
  - Sex
  - BMI
  - Day
  - Temperature
- Comorbidities:
  - Myocardial Infarction
  - Congestive Heart Failure
  - Peripheral Vascular Disease
  - Cerebrovascular Disease
  - Dementia
  - Chronic Obstructive Pulmonary Disease
  - Peptic Ulcer Disease
  - Mild Liver Disease
  - Diabetes without chronic complications
  - Diabetes with chronic complications
  - Hemiplegia or paraplegia
  - Renal disease
  - Cancer (any malignancy)
  - Metastatic solid tumor
  - Charlson score
  - Hypertension
  - Atrial fibrillation
  - Valve Replacement
  - Rheumatoid Arthritis
- Outpatient Medications taken before hospitalization (limited to medication classes taken by at least 100 patients):
  - Antihyperglycemic, Biguanide Type
  - Laxatives And Cathartics

- Vitamin D Preparations
- Calcium Channel Blocking Agents
- Proton-Pump Inhibitors
- Antihyperlipidemic-Hmgcoa Reductase Inhib (Statins)
- Beta-Adrenergic Agents, Inhaled, Short Acting
- Blood Sugar Diagnostics
- Anticonvulsants
- Analgesic/Antipyretics,Non-Salicylate
- Beta-Adrenergic Blocking Agents

• Lab Values:

  - Potassium (0.5% Missing)
  - Ferritin (8.3% Missing)
  - Calcium (0.5% Missing)
  - Neutrophil % (0.0% Missing)
  - Lymphocyte % (0.0% Missing)

Neutrophil-Lymphocyte Ratio is calculated by dividing Neutrophil % by Lymphocyte %. We exclude all patients who are missing either of these lab values.

The patient population has changed over time. The majority of patients were admitted from days 30-50. As Figure S1 shows, this period also contained the majority of patients with extremely elevated NLR levels. However, the range of NLR observed in the days and months following the initial peak of the pandemic remains wide. Many factors have shifted over the course of the pandemic, so we also control for patient intake day in our models.

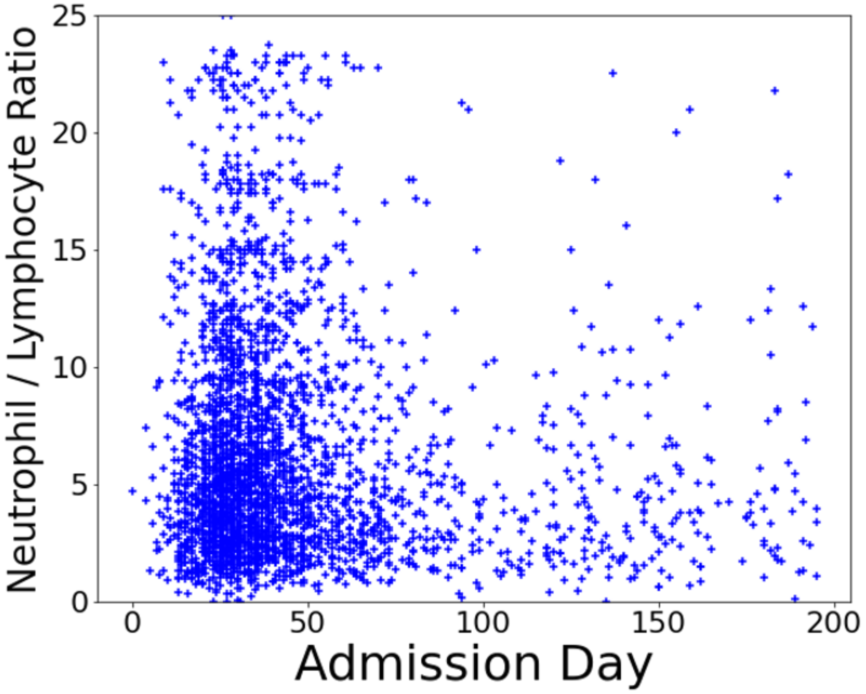


Figure S1: Distribution of patients by NLR and Admission Day. Each marker indicates a single patient, with the vertical axis indicating NLR value on the initial lab test and the horizontal axis indicating the day the patient was admitted.

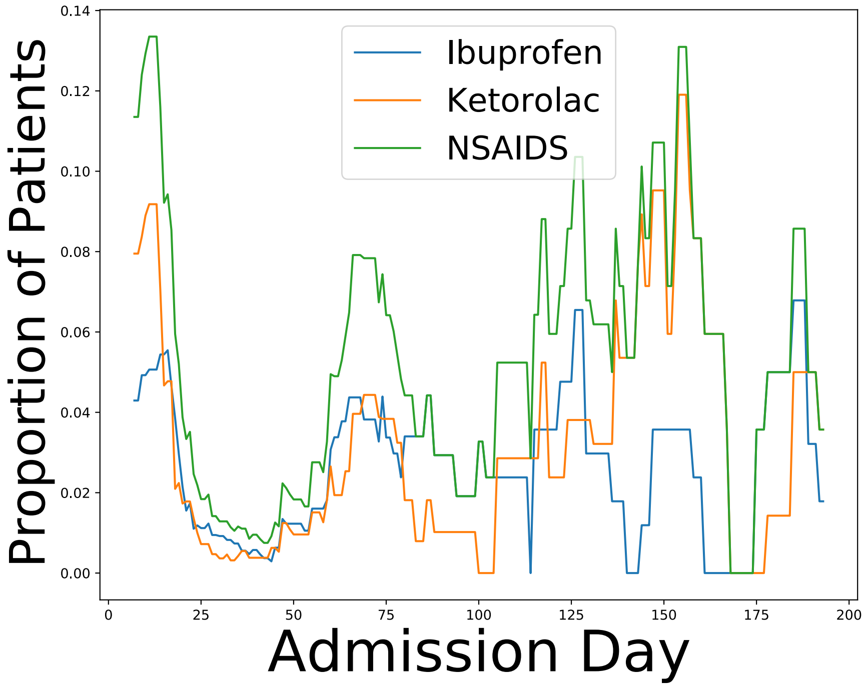


Figure S2: The proportion of patients being prescribed NSAIDs has increased over time. The values are smoothed over a 14-day period by a running average.

#### S2 Treatment and Outcome Measure Construction

In this study, we are interested in the effect of NSAIDs. To ensure a proper linking of lab values, treatments, and outcomes, we consider only NSAID treatments that are given within 24 hours of the initial lab test and at least 24 hours before mortality. Figure S2 shows that the proportion of Covid-19 patients being prescribed NSAIDs has changed over time in response to the changing guidance. Patient factors correlated and anti-correlated with NSAID treatments are shown in Tables S1,S2, respectively.

| **Patient Feature** | **R** |
|---|---|
| flag.hallucinogens | 0.046 |
| CARDIOVASCULAR DIAGNOSTICS-RADIOPAQUE | 0.046 |
| ANALGESIC/ANTIPYRETICS,NON-SALICYLATE | 0.050 |
| CEPHALOSPORIN ANTIBIOTICS - 3RD GENERATION | 0.055 |
| flag.gerd | 0.055 |
| SKELETAL MUSCLE RELAXANTS | 0.064 |
| PENICILLIN ANTIBIOTICS | 0.068 |
| OPIOID ANALGESICS | 0.075 |
| Day | 0.076 |
| INTESTINAL MOTILITY STIMULANTS | 0.083 |

Table S1: Features correlated with NSAID treamtent.

| **Patient Feature** | **R** |
|---|---|
| Age | -0.216 |
| HCQ | -0.131 |
| POTASSIUM | -0.118 |
| Charlson score | -0.114 |
| CALCIUM | -0.105 |
| Diabetes without chronic complications | -0.082 |
| Renal disease | -0.075 |
| Diabetes with chronic complications | -0.074 |
| Cerebrovascular disease | -0.073 |
| MACROLIDE ANTIBIOTICS | -0.072 |
| Congestive heart failure | -0.070 |
| flag.afib | -0.065 |

Table S2: Features anti-correlated with NSAID treatment.

As expected, NSAID usage is heavily influenced by patient age. Figure S3 shows the distribution of the ages of patients for each treatment. In-patient prescription of NSAIDs is associated with younger patients, with the effect most pronounced for Ketorolac. As expected, out-patient use of NSAIDs is assocaited with older patients.

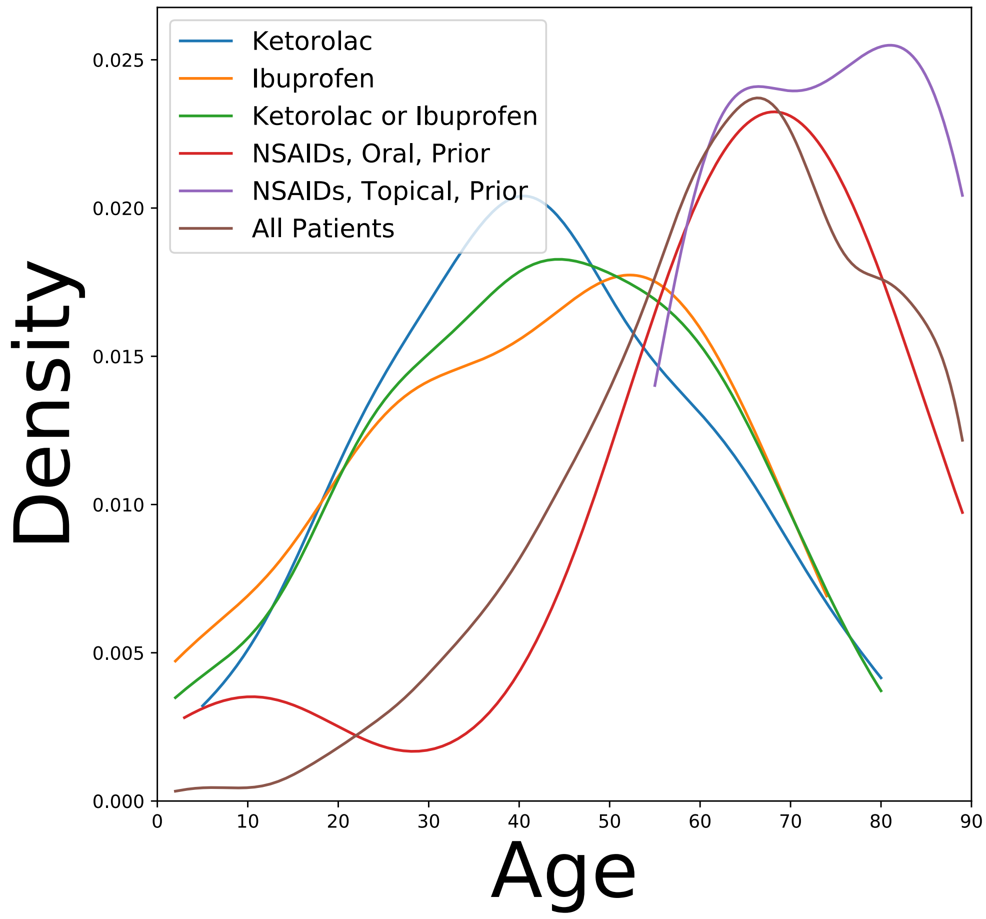


Figure S3: Distribution of ages of patients stratified by treatment.

### S3 Mortality Risk Model

We use generalized additive models (GAMs) to model patient mortality risk at admission. GAMs are a version of logistic regression that are able to model non-linear effects [12]. While logistic regression summarizes the influence of each feature with a single coefficient, GAMs estimate the influence of a feature for every value the feature can take on as a graph. This means that GAMs naturally accommodate non-linear effects, which improves both model accuracy and interpretation. Modeling non-linear effects is particularly important when features have multiple regions of high or low risk (e.g., both hyperthermia and hypothermia are associated with high risk).

We use tree-based GAMs [13] implemented in the Python `Interpret` package[2]. These GAMs are invariant to all monotonic feature transforms, so log-transforms of lab values are not necessary.

The risk model achieves an ROC of $0.912 \pm 0.001$ and an F1-score of $0.598 \pm 0.002$ on held-out patients. This significantly outperforms a logistic regression model which achieves an ROC of $0.859 \pm 0.001$ and F1-score of $0.455 \pm 0.002$ on the same data.

The 5 most important features to this background risk model are:

1. Temperature
2. Age
3. Admission Day
4. Calcium
5. Charlson score

### S4 Interpretation of Main Results

To correct for confounding, we use the mortality risk model described in S3 to correct for all risk which can be attributed to factors other than NSAIDs. We are interested in

$$P_1(x, y) = P(\text{mortality}|X = x, \text{NSAIDs} = 1),$$
$$P_0(x, y) = P(\text{mortality}|X = x, \text{NSAIDs} = 0).$$

Comparing $P_1$ and $P_0$ separates mortality risk that can only be assigned to NSAIDs treatment out from mortality risk which could be assigned to other risk factors.

We follow the convention of [11] to estimate these quantities using a classification model. We use the additive model assumption that $P(\text{mortality}|X = x, \text{NSAIDs} = z) = f(x) + g(z)$. This means that a single $f$ (the background risk GAM trained in S3) is shared between both groups of patients and that $g$ is an adjustment to this background risk model. We can estimate each $g$ as the residual of the predictions of the background risk model on a set of held-out patients. The adjusted risk ratio (ARR) is the ratio $\frac{P_1}{P_0}$, while the adjusted risk difference (ARD) is the difference $P_1 - P_0$ [10, 11]. For both ARR and ARD, lower values indicate protective effects. The ARR provides the protective effect as a relative measure while the ARD provides the absolute difference in risk between the groups.

[2] https://github.com/interpretml/interpret
